# Sociodemographic Factors Associated with Antenatal Steroid Exposure among Late Preterm Births

**DOI:** 10.1101/2024.01.22.24301562

**Authors:** Mark A. Clapp, Jessica L. Cohen, Cynthia Gyamfi-Bannerman, Anjali J. Kaimal, Scott A Lorch, Jason D. Wright, Alexander Melamed

**Author notes:** Corresponding Author: Mark A Clapp, MD MPH, Department of Obstetrics, Gynecology, and Reproductive Biology, Massachusetts General Hospital, 55 Fruit St, Boston, MA 02114. Disclosures: Dr. Clapp reports grants from the Society for Maternal-Fetal Medicine and the Risk Management Foundation of the Harvard Medical Institutions, Inc., and serves as a medical advisory board member with private equity in Delfina Health, outside the submitted work. Dr. Melamed reports grants from the National Center for Advancing Translational Sciences, the National Cancer Institute, and the Conquer Cancer-The ASCO Foundation, and the Department of Defense outside the submitted work. Dr. Melamed has also served as an advisor for AstraZeneca. Dr. Wright has received research funding from Merck and honoraria from UpToDate.

## Abstract

As the risk of preterm birth is known to vary widely, we aimed to determine if antenatal steroid exposure among LPBs varied based on sociodemographic characteristics. We hypothesized that sociodemographic factors may influence a provider ‘s clinical judgment of a patient ‘s likelihood of preterm birth and, therefore, also be associated with antenatal steroid use. This cross-sectional analysis used the 2021 US natality data and included non-anomalous, liveborn, hospital-based singleton births at ≥34 weeks of gestation to mothers without diabetes, a cohort similar to those in the ALPS Trial. The following sociodemographic factors were compared among those who received vs. did not receive steroids using chi-square tests: age, race (as designated and categorized on the birth certificate), ethnicity, primary pay for the delivery, marital status, and education. In 2021, 237,025 late preterm births met eligibility criteria, of which 17.3% were exposed to antenatal steroids. Among the following sociodemographic factors, the odds of antenatal steroid receipt were lower compared to the reference majority population: 1) Black (adjusted odds ratio (aOR) 0.88 (95% CI 0.81, 0.96)) and Native Hawaiian or Other Pacific Islander (NHOPI) (aOR 0.58 (95% CI 0.43, 0.79) compared to White race; 2) less than high school education (aOR 0.76 (95% CI 0.72, 0.81)) or high school education (Aor 0.87 (95% CI 0.83, 0.91)) compared to post-secondary education; and 3) public (aOR 0.83 (95% CI 0.78, 0.87)) or no insurance (aOR 0.68 (95% CI 0.61, 0.77)) compared to private insurance. Age and marital status were not associated with steroid receipt. Despite no known differential treatment effects of antenatal steroids by sociodemographic factors, steroid exposure among LBPs varied significantly among races, ethnicities, payers, and education levels in the US.

## Objective

In 2016, the Antenatal Late Preterm Steroid Trial demonstrated the benefit of antenatal steroids in reducing respiratory morbidity among late preterm births (“LPB,” 34-36 weeks of gestation).^1^ Prior studies have shown that this trial and its dissemination resulted in increased steroid use in the late preterm period in the US, though adoption was not uniform.^2–4^ As the risk of preterm birth is known to vary widely, we aimed to determine if antenatal steroid exposure among LPBs varied based on sociodemographic characteristics.^5^ We hypothesized that sociodemographic factors may influence a provider ‘s clinical judgment of a patient ‘s likelihood of preterm birth and, therefore, also be associated with antenatal steroid use.

## Study Design

This cross-sectional analysis used the 2021 US natality data and included non-anomalous, liveborn, hospital-based singleton births at ≥34 weeks of gestation to mothers without diabetes, a cohort similar to those in the ALPS Trial.^1^ The primary outcome was the receipt of steroids, as reported on the birth certificate. The following sociodemographic factors were compared among those who received vs. did not receive steroids using chi-square tests: age, race (as designated and categorized on the birth certificate), ethnicity, primary pay for the delivery, marital status, and education. A multivariable logistic regression model was constructed with all the sociodemographic and two clinical factors hypothesized to influence clinical decision-making (parity and week of gestation at delivery). Models used cluster robust standard errors based on county of birth. Missing data were included in the model as “unknown.” P-values <0.05 were considered statistically significant. The Mass General Brigham institutional review board categorized this as non-human subjects research.

## Results

In 2021, 237,025 late preterm births met eligibility criteria, of which 17.3% were exposed to antenatal steroids. Among the following sociodemographic factors, the odds of antenatal steroid receipt were lower compared to the reference majority population (Table 1): 1) Black race (adjusted odds ratio (aOR) 0.88 (95% CI 0.81, 0.96)) and Native Hawaiian or Other Pacific Islander (NHOPI) (aOR 0.58 (95% CI 0.43, 0.79) compared to White race; 2) less than high school education (aOR 0.76 (95% CI 0.72, 0.81)) or high school education (aOR 0.87 (95% CI 0.83, 0.91)) compared to post-secondary education; and 3) Medicaid (aOR 0.83 (95% CI 0.78,0.87)) or no insurance (aOR 0.68 (95% CI 0.61, 0.77)) compared to private insurance. Age and marital status were not associated with steroid receipt. Of the two clinical factors included, nulliparity and earlier gestational ages were associated with increased odds of steroid receipt.

**Table 1:**
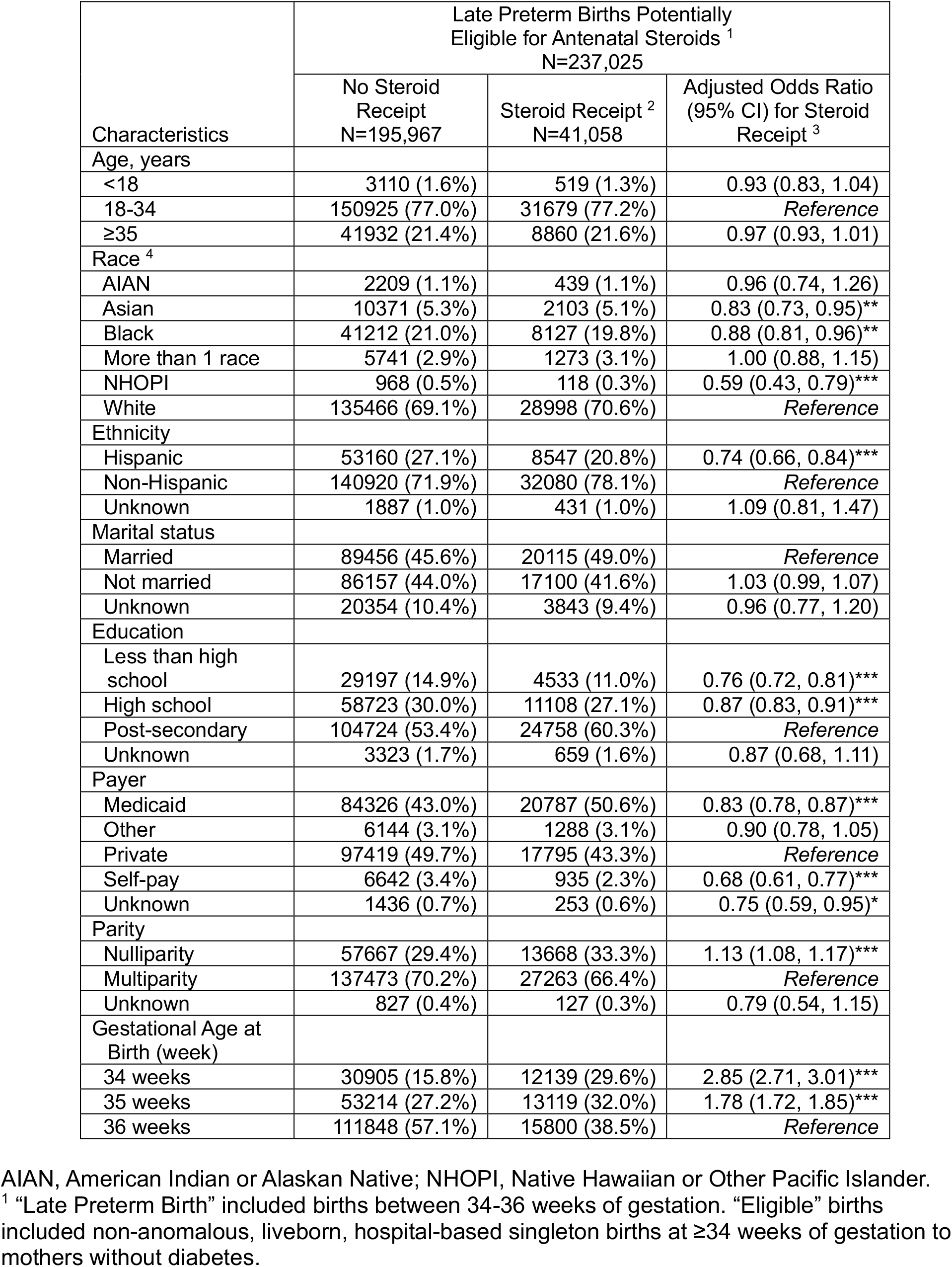
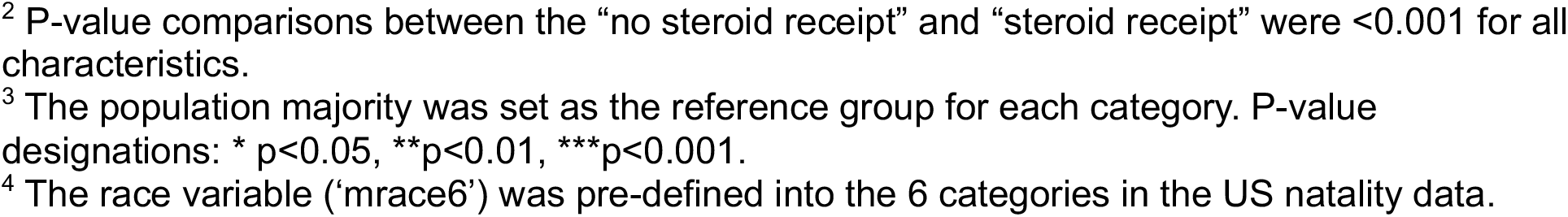
Antenatal Steroid Exposure by Sociodemographic Factors.

| Characteristics | Late Preterm Births Potentially Eligible for Antenatal Steroids <sup>1</sup><br>N=237,025 |  |  |
| --- | --- | --- | --- |
|  | No Steroid Receipt<br>N=195,967 | Steroid Receipt <sup>2</sup><br>N=41,058 | Adjusted Odds Ratio (95% CI) for Steroid Receipt <sup>3</sup> |
| Age, years |  |  |  |
| <18 | 3110 (1.6%) | 519 (1.3%) | 0.93 (0.83, 1.04) |
| 18-34 | 150925 (77.0%) | 31679 (77.2%) | Reference |
| ≥35 | 41932 (21.4%) | 8860 (21.6%) | 0.97 (0.93, 1.01) |
| Race <sup>4</sup> |  |  |  |
| AIAN | 2209 (1.1%) | 439 (1.1%) | 0.96 (0.74, 1.26) |
| Asian | 10371 (5.3%) | 2103 (5.1%) | 0.83 (0.73, 0.95)** |
| Black | 41212 (21.0%) | 8127 (19.8%) | 0.88 (0.81, 0.96)** |
| More than 1 race | 5741 (2.9%) | 1273 (3.1%) | 1.00 (0.88, 1.15) |
| NHOPI | 968 (0.5%) | 118 (0.3%) | 0.59 (0.43, 0.79)*** |
| White | 135466 (69.1%) | 28998 (70.6%) | Reference |
| Ethnicity |  |  |  |
| Hispanic | 53160 (27.1%) | 8547 (20.8%) | 0.74 (0.66, 0.84)*** |
| Non-Hispanic | 140920 (71.9%) | 32080 (78.1%) | Reference |
| Unknown | 1887 (1.0%) | 431 (1.0%) | 1.09 (0.81, 1.47) |
| Marital status |  |  |  |
| Married | 89456 (45.6%) | 20115 (49.0%) | Reference |
| Not married | 86157 (44.0%) | 17100 (41.6%) | 1.03 (0.99, 1.07) |
| Unknown | 20354 (10.4%) | 3843 (9.4%) | 0.96 (0.77, 1.20) |
| Education |  |  |  |
| Less than high school | 29197 (14.9%) | 4533 (11.0%) | 0.76 (0.72, 0.81)*** |
| High school | 58723 (30.0%) | 11108 (27.1%) | 0.87 (0.83, 0.91)*** |
| Post-secondary | 104724 (53.4%) | 24758 (60.3%) | Reference |
| Unknown | 3323 (1.7%) | 659 (1.6%) | 0.87 (0.68, 1.11) |
| Payer |  |  |  |
| Medicaid | 84326 (43.0%) | 20787 (50.6%) | 0.83 (0.78, 0.87)*** |
| Other | 6144 (3.1%) | 1288 (3.1%) | 0.90 (0.78, 1.05) |
| Private | 97419 (49.7%) | 17795 (43.3%) | Reference |
| Self-pay | 6642 (3.4%) | 935 (2.3%) | 0.68 (0.61, 0.77)*** |
| Unknown | 1436 (0.7%) | 253 (0.6%) | 0.75 (0.59, 0.95)* |
| Parity |  |  |  |
| Nulliparity | 57667 (29.4%) | 13668 (33.3%) | 1.13 (1.08, 1.17)*** |
| Multiparity | 137473 (70.2%) | 27263 (66.4%) | Reference |
| Unknown | 827 (0.4%) | 127 (0.3%) | 0.79 (0.54, 1.15) |
| Gestational Age at Birth (week) |  |  |  |
| 34 weeks | 30905 (15.8%) | 12139 (29.6%) | 2.85 (2.71, 3.01)*** |
| 35 weeks | 53214 (27.2%) | 13119 (32.0%) | 1.78 (1.72, 1.85)*** |
| 36 weeks | 111848 (57.1%) | 15800 (38.5%) | Reference |

AIAN, American Indian or Alaskan Native; NHOPI, Native Hawaiian or Other Pacific Islander.
<sup>1</sup> “Late Preterm Birth” included births between 34-36 weeks of gestation. “Eligible” births included non-anomalous, liveborn, hospital-based singleton births at ≥34 weeks of gestation to mothers without diabetes.
<sup>2</sup> P-value comparisons between the “no steroid receipt” and “steroid receipt” were <0.001 for all characteristics.
<sup>3</sup> The population majority was set as the reference group for each category. P-value designations: \* $p < 0.05$ , \*\* $p < 0.01$ , \*\*\* $p < 0.001$ .
<sup>4</sup> The race variable ('mrace6') was pre-defined into the 6 categories in the US natality data.

## Conclusion

Despite no known differential treatment effects of antenatal steroids by sociodemographic factors, steroid exposure among LBPs varied significantly among races, ethnicities, payers, and education levels in the US. Many of the same subgroups with lower odds of steroid receipt have the highest preterm birth rates.^5^ There are several possible explanations, including 1) provider assessment of baseline risk of preterm delivery does not influence the likelihood of steroid receipt, 2) underlying biases affect clinical judgment that results in disparate use; 3) patients have a varying willingness to receive intervention; or 4) differential clinical and structural (e.g., distance to hospital) factors influence the opportunity to intervene (e.g., patients presenting “too late” to receive the intervention). The study used US birth certificate data; thus, it was limited in its ability to observe the exact timing of steroid administration or other clinical factors that may have affected clinical decision-making. Additional studies examining the drivers of these sociodemographic differences, such as patient perception of risk, clinician judgment, and hospital or regional factors affecting steroid use, are needed to inform policy solutions to address disparities in antenatal steroid use, especially among populations at the highest risk for preterm birth.

## Data Availability

All data used in this analysis are available online at cdc.gov.

## Acknowledgments

The authors have no acknowledgments.

## References

1. Gyamfi-Bannerman C, Thom EA, Blackwell SC, et al. Antenatal Betamethasone for Women at Risk for Late Preterm Delivery. New England Journal of Medicine. 2016;374(14):1311–1320. doi:10.1056/NEJMoa1516783

2. Clapp MA, Melamed A, Freret TS, James KE, Gyamfi-Bannerman C, Kaimal AJ. US Incidence of Late-Preterm Steroid Use and Associated Neonatal Respiratory Morbidity After Publication of the Antenatal Late Preterm Steroids Trial, 2015-2017. JAMA Netw Open. 2022;5(5):e2212702. doi:10.1001/jamanetworkopen.2022.12702

3. Freret TS, Cohen JL, Gyamfi-Bannerman C, et al. Regional Variation in Antenatal Late Preterm Steroid Use following the ALPS Trial. medRxiv. 2023;2023.05.25.23290522. 10.1101/2023.05.25.23290522

4. Society for Maternal-Fetal Medicine (SMFM) Publications Committee. Implementation of the use of antenatal corticosteroids in the late preterm birth period in women at risk for preterm delivery. Am J Obstet Gynecol. 2016;215(2):B13–15. doi:10.1016/j.ajog.2016.03.013

5. Manuck TA. Racial and ethnic differences in preterm birth: A complex, multifactorial problem. Semin Perinatol. 2017;41(8):511–518. doi:10.1053/j.semperi.2017.08.010

